# Measure by Measure: Resting Heart Rate Across the 24 Hour Cycle

**DOI:** 10.1101/2021.11.24.21266320

**Authors:** CA Speed, TC Arneil, RK Harle, A Wilson, A Karthikesalingam, MV McConnell, JP Phillips

## Abstract

**Background:** Photoplethysmography (PPG) sensors, typically found in wrist-worn devices, can continuously monitor heart rate (HR) in large populations in real-world settings. Resting heart rate (RHR) is an important biomarker of morbidities and mortality, but no universally accepted definition nor measurement criteria exist. In this study, we provide a working definition of RHR and describe a method for accurate measurement of this biomarker, recorded using PPG derived from wristband measurement across a 24 hour cycle.

**Methods:** 433 healthy subjects wore a wrist device that measured activity and HR for up to 3 months. HR during inactivity was recorded and the duration of inactivity needed for HR to stabilise was ascertained. We identified the lowest HR during each 24-hour cycle (true RHR) and examined the time of day or night this occurred. The variation of HR during inactivity through the 24-hour cycle was also assessed. The sample was also subdivided according to daily activity levels for subset analysis.

**Findings:** Adequate data was obtained for 19,242 days and 18,520 nights. HR stabilised in most subjects after 4 minutes of inactivity. Mean (SD) RHR for the sample was 54.5 (8.0) bpm (day) and 50.5 (7.6) bpm (night). RHR values were highest in the least active group (lowest MET quartile). A circadian variation of HR during inactivity was confirmed, with the lowest values being between 0300 and 0700 hours for most subjects.

**Interpretation:** RHR measured using a PPG-based wrist-worn device is significantly lower at night than in the day, and a circadian rhythm of HR during inactivity was confirmed. Since RHR is such an important health metric, clarity on the definition and measurement methodology used is important. A minimum rest time of 4 minutes provides a reliable measurement of HR during inactivity and true RHR in a 24-hour cycle is best measured between 0300 and 0700 hours.

**Funding:** This study was funded by Google.

Research In context
Resting heart rate is one of the most fundamental and relevant biomarkers in health and disease. Its measurement, particularly in large population studies, is increasingly reliant on sensor devices. No universally accepted definition nor measurement criteria exist. In this study, we provide a working definition of RHR and describe a method for measurement of RHR recorded using PPG derived from wristband measurement. We make recommendations to support future studies of resting heart rate using wearable sensors.

## Introduction

Resting heart rate (RHR) is an important biomarker of health and disease [1-7]. It is a vital sign, an indicator of all cause mortality [1,2,7], cardiometabolic risk [1-9], cardiorespiratory fitness [10] and is used in regular monitoring to indicate general response to training [10] and is a highly variable physiological marker with many influencing factors. There is considerable inconsistency among definitions and methodological criteria for assessment of RHR, and consumer devices that provide estimations of RHR to their users show considerable variability in methodology and reported values. The term “resting heart rate” sometimes is used to represent the heart rate (HR) of an individual whenever they are inactive, as opposed to the lowest HR at rest in a 24-hour cycle. Methodological variables including time of measurement, subject positioning, pre-measurement rest, and mode of measurement (pulse palpation, ECG, and photoplethysmography (PPG) measured using wearable technologies such as a wristband or watch) may all influence the measurement. With the increasing availability of continuous HR monitoring using wearable devices, and the expansion of use of these devices in health monitoring, clarity on definitions and the differences between measurements is important, and within reach.

PPG-sensor technology has been demonstrated to accurately measure HR when compared to ECG and is increasingly used in cardiovascular health studies [11, 12]. Note that PPG devices vary in their design and algorithms used to calculate HR in activity and at rest. The use of PPG-based wearable devices offers new opportunities in research across large populations. In a retrospective study of over 92,000 subjects wearing a range of different wrist-worn devices, mean ‘RHR’ was reported to range from 40-109 bpm across a US-based population with a mean (SD) age 45.8 (14.4) years [15]. RHR was provided using a proprietary formula from the device maker, with a single value being provided for each day, but no further insights were provided as to the methodology of its estimation. To our knowledge, no other PPG-based study has provided information as to methodology of RHR calculation.

In this real-world study, our primary aims were to describe a method for measurement of RHR recorded using PPG derived from wrist-worn fitness tracker measurements, to demonstrate the potential for erroneous estimation of RHR, and to assess quantitatively how long it takes HR to stabilise after activity ceases. We also evaluated the time of day on HR when inactive, examining for the presence of a circadian rhythm of HR during inactivity across 24 hours, and we evaluated the timing of RHR across subjects. We also further scrutinised the lowest HR during inactivity in the day compared to values at night.

## Methods

### Study population

The study targeted healthy adults living within the United States aged 18-65 years. Eligible participants were physically active, as assessed by the International Physical Activity Questionnaire (IPAQ) [16, 17]. The study was approved by an Institutional Review Board (Advarra, Columbia, MD). Consenting individuals agreed to wear a wrist device, the Fitbit Charge 4 fitness tracker (Fitbit LLC, San Francisco, CA, USA), continuously for up to three months. The study population comprised 433 subjects (66.1% female), mean(+-SD) age = 35.4 +- 9.6 years. Mean (+-SD) BMI was 26.5 (+- 5.2). No subjects had known cardiovascular disease and none was on antiarrhythmics. Subjects with a pacemaker and those who were night shift workers were excluded.

### Data collection

The wrist-worn device captured HR using PPG sensors and activity through accelerometry. Participants were asked to perform their normal daily and night-time routines while wearing the device as often as possible. The device charges rapidly and subjects were asked to charge the device whilst bathing. Data from each device was uploaded automatically to a central secure platform (the Fitabase research platform [18]).

HR and estimates of activity level from accelerometry were recorded by the device every 60 seconds. HR during inactivity across the day and night was derived from these HR by taking the lowest HR measured by the device when still, i.e. when the metabolic equivalent (MET) during each 60 second epoch was estimated to be less than 1.5. The device itself has filtering algorithms to remove noise, although it should be noted that when subjects are inactive, the HR accuracy is typically reliable.

Chow et al. found that the mean absolute percentage error (MAPE) of HR measured using fitness trackers in young adults at rest compared to an ECG chest strap (Polar M3) was 3.96% and 4.46% for the Garmin Vivosmart HR+ and Xiaomi Mi Band 2 devices respectively [19].

The wrist-worn device has built-in algorithms to determine the start and end times that the wearer was in bed, which were logged to enable separate calculations of lowest HR during inactivity in the day as well as at night. This implicitly defined night-time for a user as the primary sleep period identified by the tracker’s algorithms.

### Data analysis

Time series of HR and physical activity were analysed to produce estimates of daytime and night-time HR during inactivity. The RHR for each day and each night were determined by taking the lowest HR recorded when the wearer was inactive during the day and at night, respectively. Night-time and daytime heart rates were distinguished from automated logging of bed start and end times by the Fitbit device. For daytime calculations, the device must have been worn for at least 8 hours in the daytime and for night-time calculations, for at least 3 hours at night.

Activity recognition on the device allowed sedentary (MET < 1.5) times to be identified, and HR during those periods was scrutinised. The time taken for the HR to stabilise during inactivity was examined to identify the necessary duration of rest needed to allow best estimate of the RHR value.

HR over the daytime and night-time were compared and the variation of HR during inactivity across the 24-hour cycle was investigated. We also examined the effects of average activity levels on RHR.

### Findings

Of 26,758 possible subject days and nights in the study, a total of 19,242 days of daytime data (>8 hours of use), and 18,520 nights (>3 hours of sleep) of data were obtained. This represents 71.9% compliance for daytime wear, and 69.2% compliance for night-time use. Of the qualifying data, the mean (SD) hours worn in a 24hr period was 15.19 (2.01) hours and at night was 7.97 (1.41) hours. The night-time mean start (bedtime) and finish (rise) times were 23:02 and 06:27, respectively.

To identify the minimum acceptable time for the HR to stabilise, periods of inactivity of 20 minutes or more were identified. This identified 160,357 sections of data (day) and 44,502 (night). The number of sampling sections was lower at night as the duration of each sampling period was longer when the subjects were consistently inactive when asleep. The HR recorded during each minute was compared with the HR from the preceding minute. The HR differences for adjacent minutes for the first 7 minutes during day and night are shown in Table One. The mean decrease in HR between 3 and 4 minutes after a period of inactivity begins is less than 0.5 bpm. After 4 minutes of inactivity, a further mean decrease in HR of less than 1.0 bpm was observed when compared to 20 minutes of inactivity. The mean HR values recorded during each inactive minute are shown in Figure One.

**Table One:** Change in heart rate during inactivity.

|  | **DAYTIME (*n*=160,357)** | **NIGHTTIME (*n*=44,502)** |
| --- | --- | --- |
| **Adjacent inactive mins** | **Mean decrease in HR from preceding minute (bpm)** | **Mean decrease in HR from preceding minute (bpm)** |
| 0-1 | 2.99 | 1.63 |
| 1-2 | 3.22 | 2.21 |
| 2-3 | 0.96 | 0.41 |
| 3-4 | 0.42 | 0.24 |
| 4-5 | 0.24 | 0.14 |
| 5-6 | 0.17 | 0.06 |
| 6-7 | 0.16 | 0.03 |

**Figure One:**
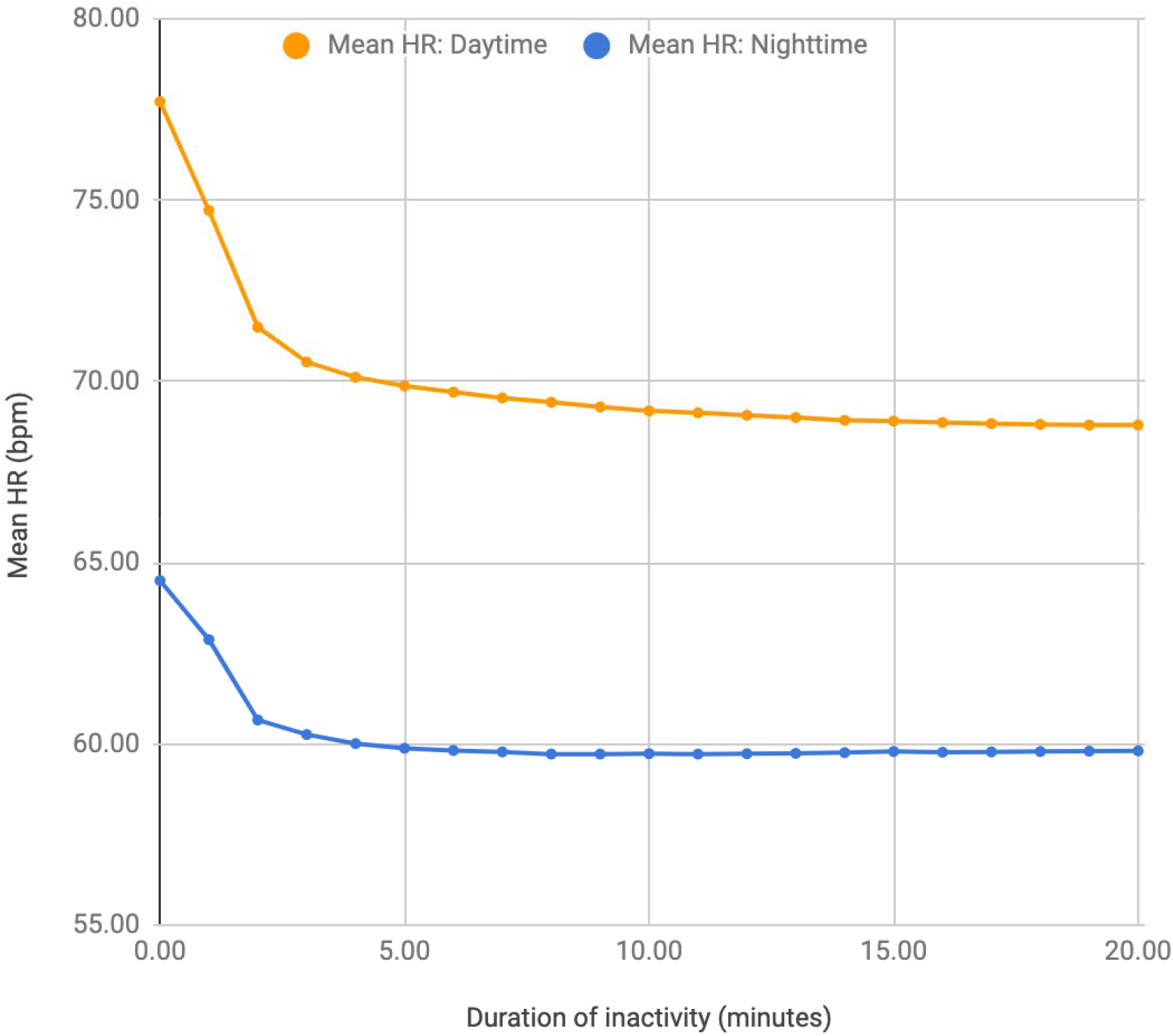
Decline in mean heart rate after initiation of inactivity.

There are many examples in the data where the HR took longer to stabilise. This phenomenon was investigated by examining the percentage of the sample where HR during inactivity fell by more than 1, 2 or 3 beats per minute in each time interval. The results of this analysis are shown in Table Two. The HR in over 13% of subjects had not stabilised, i.e., the change in HR from 14 to 15 minutes of inactivity is greater than 3 bpm.

**Table Two:** Percentage of sample where heart rate during inactivity had not stabilised.

| **Adjacent inactive mins** | **% of sample where**<br>**change in HR > 1 bpm** | **% of sample where**<br>**change in HR > 2 bpm** | **% of sample where**<br>**change in HR > 3 bpm** |
| --- | --- | --- | --- |
| **DAYTIME** |  |  |  |
| 0-1 | 54.9 | 46.9 | 39.9 |
| 1-2 | 61.3 | 51.5 | 42.7 |
| 2-3 | 44.2 | 33.0 | 24.4 |
| 3-4 | 38.7 | 27.0 | 18.6 |
| 14-15 | 33.0 | 21.1 | 13.1 |
| **NIGHTTIME** |  |  |  |
| 0-1 | 48.9 | 40.8 | 34.3 |
| 1-2 | 52.3 | 42.0 | 34.3 |
| 2-3 | 35.9 | 23.4 | 15.9 |
| 3-4 | 32.8 | 19.1 | 11.7 |
| 14-15 | 23.8 | 12.7 | 7.6 |

RHR in a 24-hour period was taken to be the lowest HR recorded during day or night after at least four minutes of inactivity. We also compared values of the lowest HR during inactivity in the day with that at night. Night-time values were lower than those in daytime, as shown in Table Three. Note the low level of 34 bpm was measured in one very fit subject on one occasion; the mean RHR for this subject during the study was 38 bpm. The mean (SD) difference (day-night) was + 3.9 (3.8) bpm. A paired Student’s t-test showed significant differences between day and night-time RHR measurements (*t*-statistic = 136.99, *P* < 0.0001). The distribution of true RHR is illustrated in Figure Two.

**Table Three:** Summary of daytime and night-time lowest heart rates during inactivity.

|  | *N* | Mean(+SD) Heart rate | Heart rate Range |
| --- | --- | --- | --- |
| **Daytime** | 19242 | 54.5 (8.0) | 34.0 - 92.4 |
| **Nighttime** | 18520 | 50.5 (7.6) | 32.6 - 89.8 |

**Figure Two:**
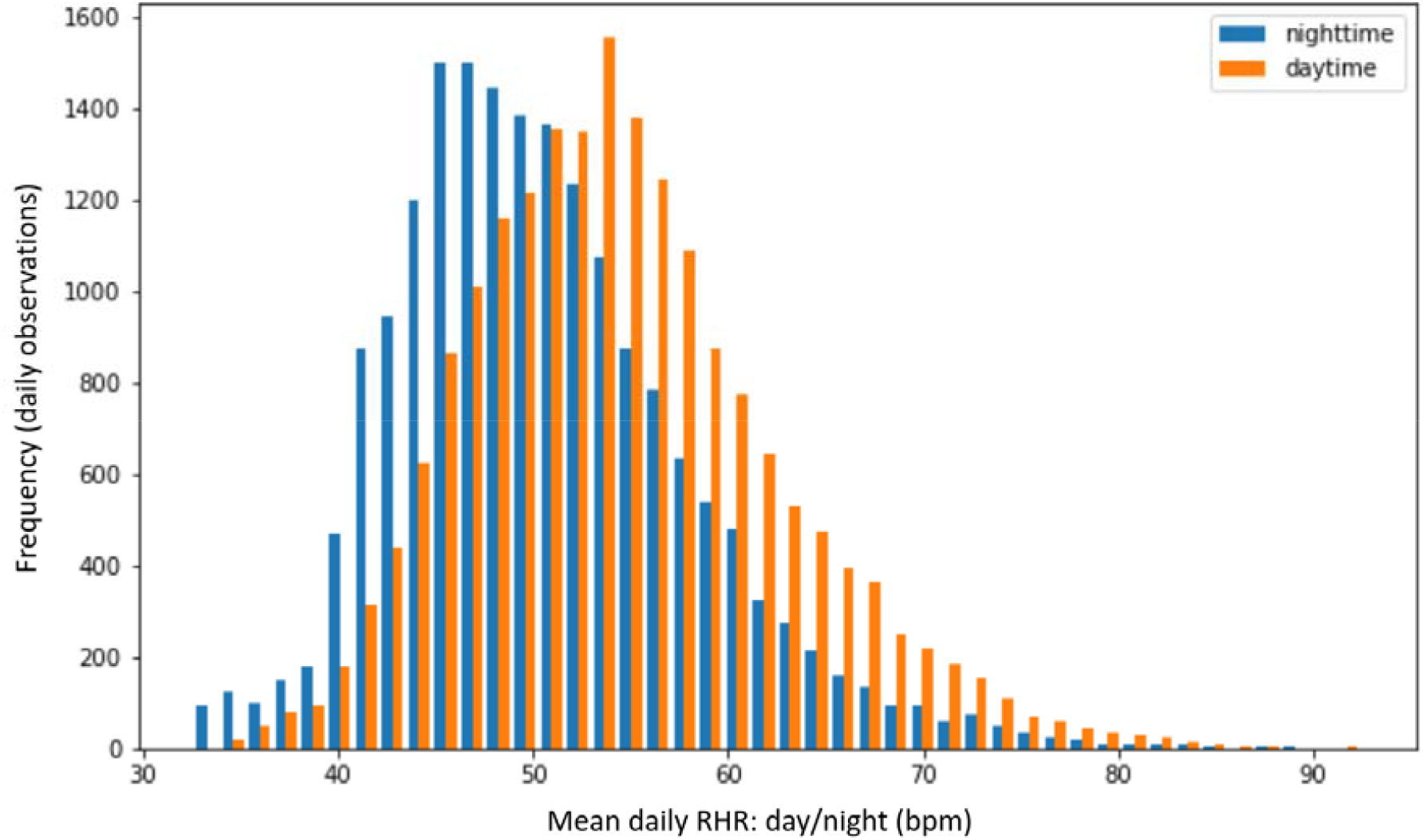
The distribution of mean RHR estimates for both daytime and night-time.

The daytime lowest HR during inactivity obtained using our method was compared with the daytime ‘RHR’ calculated using the only other available definition, i.e.: the lowest HR during the day irrespective of activity [15]. The latter ‘RHR’ mean (SD) was 53.9 (9.9), i.e. 0.6 bpm lower than our estimate, due to a small number of instances where the lowest daily HR occurs when the subject is deemed to be moving by the device. The night-time lowest HR estimates differed by less than 0.1 bpm.

We also examined the time of day that lowest HR was noted. Just over half (53%) occurred between 0300 and 0700. The distribution is shown in the histogram below (Figure Three).

**Figure Three:**
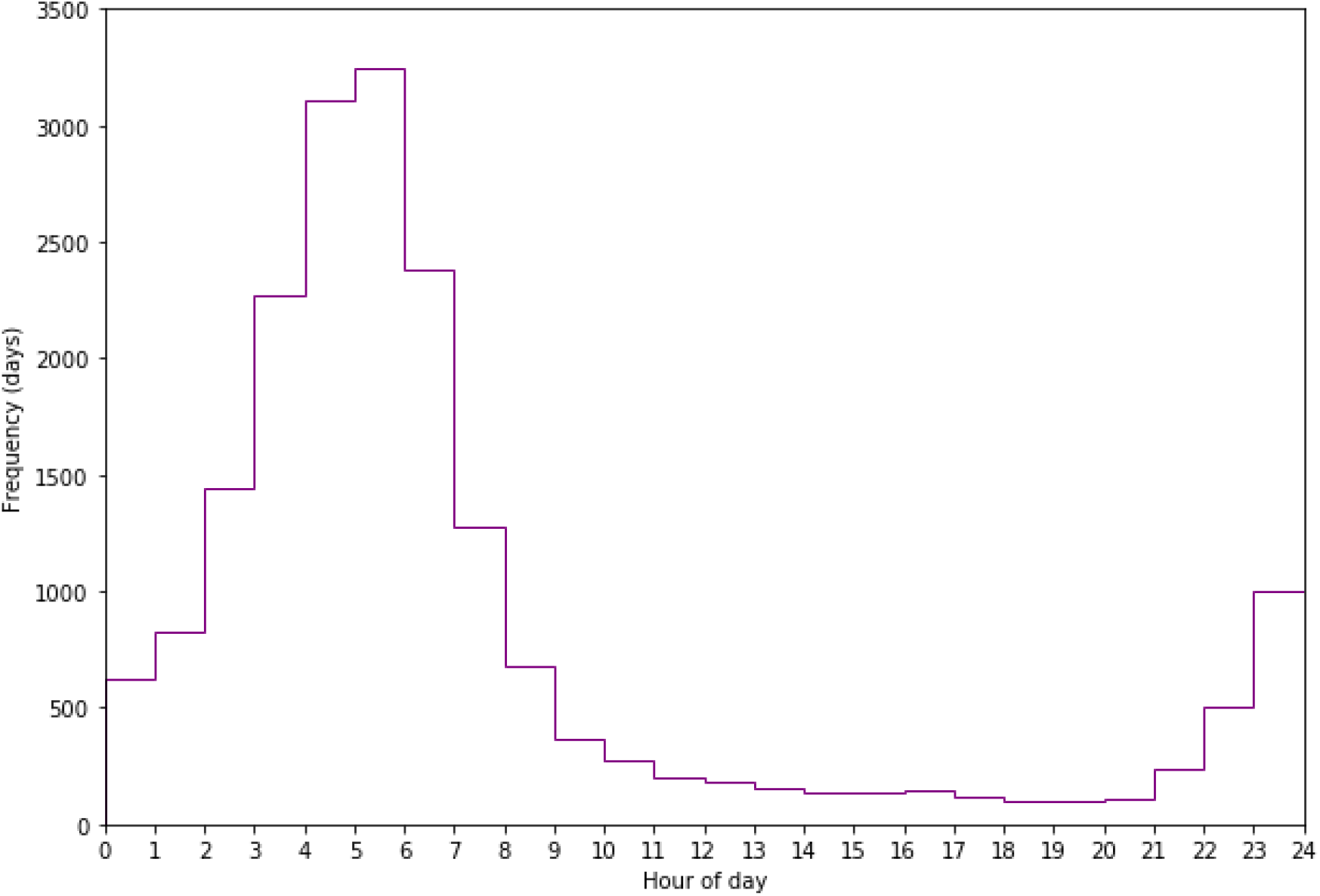
Distribution across subjects of timing of lowest RHR in 24-hour cycle, showing that on most days the RHR was lowest between 0300 and 0700 across the population.

We examined the trend in mean HR during inactivity across the 24-hour cycle during the study period (Figure Four). All HR measurements after at least 4 minutes of inactivity were averaged for all subjects for each hour of the day as indicated by the devices’ timestamps.

**Figure Four:**
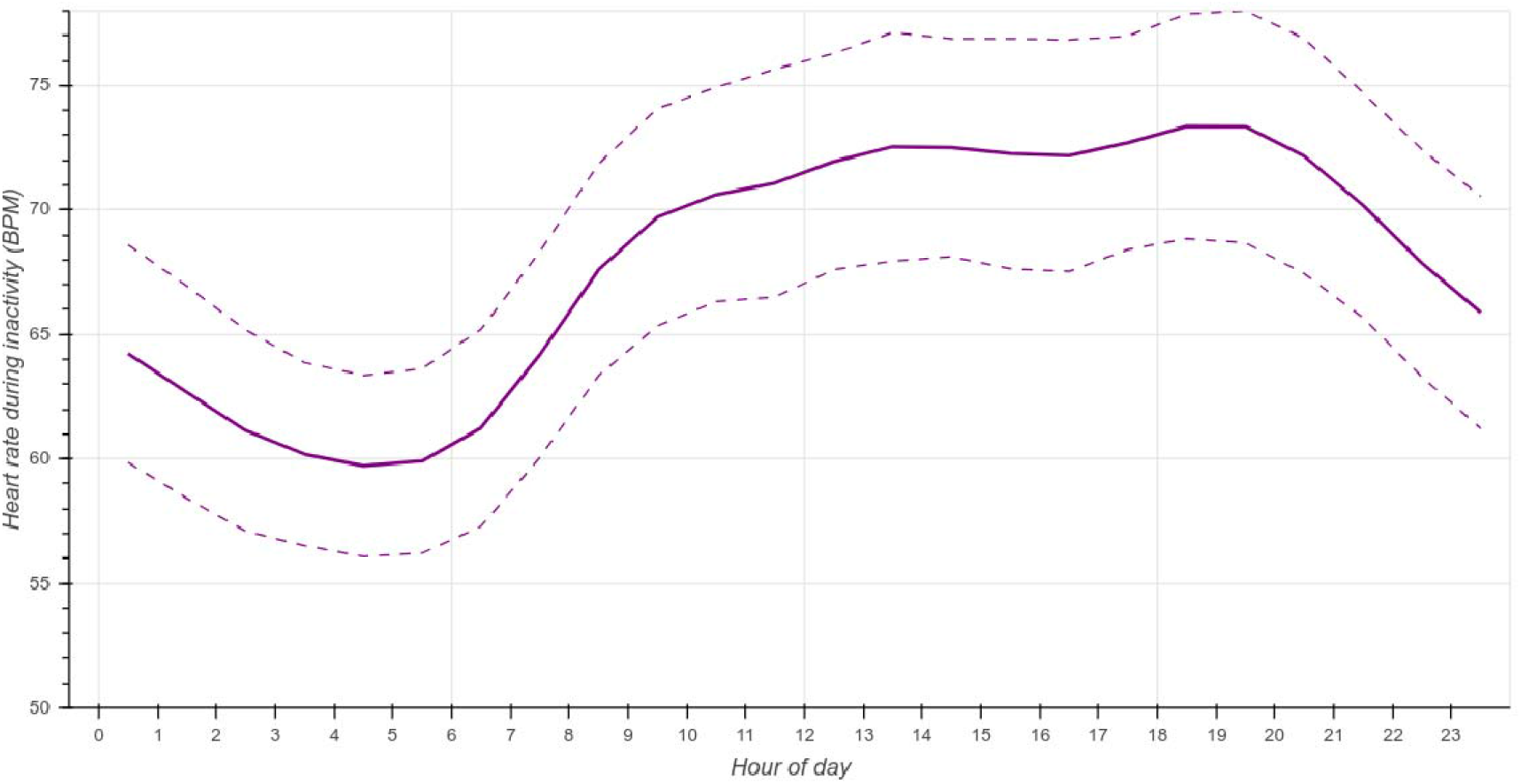
Circadian variation in heart rate during inactivity. Mean (SD) heart rate after 4 minutes of inactivity by hour of the day.

Lastly, we examined the effects of individuals’ activity levels on their lowest HR during inactivity (Table Four). The activity level for each subject was represented by the mean number of minutes of the day where the MET, estimated from the device’s accelerometer, was greater than 3.0, equivalent to standard definitions of moderate to vigorous activity (MVPA). The results were grouped by activity level quartiles (Figure Five).

**Figure Five:**
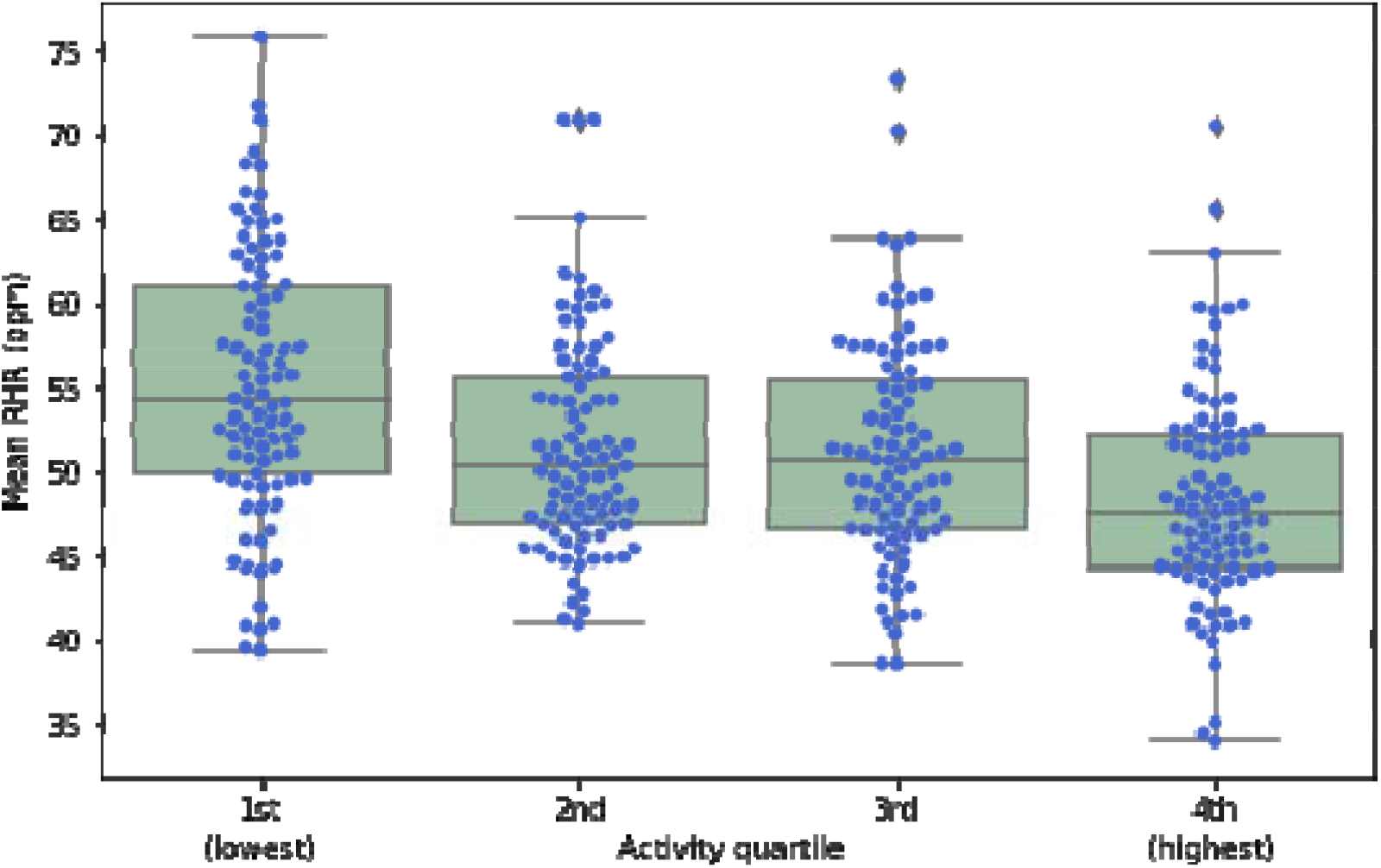
Box plots of the mean RHR in the 24-hour cycle for each activity quartile.

**Table Four:** Multiple Comparison of Means - Tukey HSD (FWER = 0.05)

| Group (1) | Group (2) | Mean diff (bpm) | p-adj | Lower | Upper | Reject? |
| --- | --- | --- | --- | --- | --- | --- |
| 1st | 2nd | -3.7005 | 0.0019 | -6.3432 | -1.0578 | True |
| 1st | 3rd | -4.0132 | 0.001 | -6.6559 | -1.3705 | True |
| 1st | 4th | -6.7866 | 0.001 | -9.4293 | -4.1439 | True |
| 2nd | 3rd | -0.3127 | 0.900 | -2.9554 | 2.330 | False |
| 2nd | 4th | -3.0861 | 0.0146 | -5.7288 | -0.4434 | True |
| 3rd | 4th | -2.7734 | 0.0355 | -5.4161 | -0.1307 | True |

Between-group differences in RHR were tested for significance using a single-factor ANOVA analysis with post-hoc subgroup comparisons. Table Four shows the results from a multiple comparison of means using Tukey’s honestly significant difference (HSD) test with a family-wise error rate (FWER), i.e. the probability of a Type I error, set to 0.05. The lowest activity and highest activity quartiles had mean (SD) RHRs of 55.2(7.94) bpm and 48.5(6.51) bpm respectively. We noted that those in the first activity quartile (i.e., individuals with lowest mean activity levels) had significantly higher RHR’s compared to those in the other three quartiles.

### Interpretation

Several studies have demonstrated an association between RHR and cardiovascular mortality both in general populations [1–3] and in patients with hypertension [4,5] or cardiac diseases [6,7]. ‘RHR’ in the studies was taken as heart rate when inactive. HR during inactivity in this context has predominantly been studied during the day [1-7] and has variably been defined as a single measure on ECG [1], or pulse palpation [2, 5, 7]. Night-time HR has been much less commonly studied but has been reported to be a better predictor of cardiovascular events than day-time lowest HR [20,21]. If the collective conclusion from these studies is that the lowest HR during inactivity (true ‘resting heart rate’) in a 24-hour cycle is the best predictor, then our study findings suggest that it is reasonable to recommend that night-time is the best time to assess this.

It is notable that the HR in some subjects took some minutes to stabilise, and indeed in over 13% of subjects it had not fully stabilised (i.e., it was decreasing at a rate greater than 3 bpm) after 15 minutes of inactivity. These results were considered most likely to have occurred due to the physiological effect of more intense exercise before the rest period. This has relevance for clinical practice, and studies where “resting heart rate” is taken by pulse after only a few minutes of rest.

The use of PPG in the monitoring of HR allows large-scale, real-world population studies that are not feasible using ECG. The increasing quality of PPG sensors has resulted in excellent quality of HR measurement in newer devices such as that used in this study. At rest, their accuracy is similar to ECG [11-14], and the inclusion of activity recognition in our study allowed us to ascertain when the subject was inactive. This is important, as when there is error in signal detection with wrist-worn devices, this is more likely when the subject is active and erroneously low HR during activity can occur.

‘Ambulatory’ HR has also been proposed to be relevant, and also has varying definitions, from the mean derived from 24-hour continuous monitoring, or from ‘snapshots’ measured during the day. It too has also been reported to have predictive value in mortality risk in elderly hypertensives [22]. Whilst it is possible to measure this using PPG, there is more likely to be error due to activity affecting signal integrity, and the capacity to measure true RHR may make ‘ambulatory’ recordings unnecessary for risk assessment.

As anticipated, RHR in our study was noted to be significantly lower in those subjects who were routinely more active, but a ‘little goes a long way’, with the most significant difference in RHR being seen between the least active and the next quartile of activity level.

We noted a circadian rhythm of HR during inactivity, and this has been studied by a few groups, who have also noted the trend we identified in our study, specifically with lower HR during the night and in the first few hours in the morning [23, 24]. This is considered to be related to the dominance of sympathetic tone and circulating catecholamines during the day, and its relative decline at night, accompanied by increased parasympathetic tone during those night-time hours [23]. Activity between rest intervals may also influence the values across the day, and this may have influenced our finding that in some samples the HR can take longer than the typical 4 minutes to fully stabilise.

It can be proposed from this data that true RHRs occur most commonly between 0300 and 0700 in the general population, apart from those who are night shift workers, who were excluded from this study.

A limitation of this study is that it may not be wholly generalizable to other wearable devices. Our estimates of HR were dependent on the Fitbit Charge 4’s segmentation of day/night and moving/sedentary periods. These factors may introduce some bias in our recommended approach to measuring RHR, so we suggest a comparison of results from a range of wearable devices as an area for future research.

## Conclusions

Resting heart rate is a vital physiological marker that reflects health, disease and mortality risk. Nevertheless, specific guidance as to how to measure it reliably is lacking. Before the availability of reliable wrist-worn devices using PPG signals, measurement has relied largely on pulse palpation and ECG-based measures. The use of PPG-based devices such as those used in our study allows specific criteria to be set relating to RHR measurement across all settings, and greater insights into its circadian variation. We recommend a minimum inactivity period of 4 minutes for reliable HR measurement to calculate RHR and suggest that subjects have also not engaged in significant exercise in the immediately preceding period. We also recommend that the true RHR in a 24-hour cycle is best measured between 0300 and 0700 hours. We encourage authors to clarify their methodology of RHR measurement in future studies.

## Data Availability

All data produced in the current work are presented in the manuscript

## REFERENCES

1. Kannel WB, Kannel C, Paffenbarger RS Jr, Cupples LA. Heart rate and cardiovascular mortality: The Framingham Study. Am Heart J. 1987; 113:1489–1494.

2. Palatini P, Casiglia E, Julius S, Pessina AC. High heart rate: a risk factor for cardiovascular death in elderly men. Arch Intern Med. 1999; 159:585–592.

3. Jouven X, Empana JP, Schwartz PJ, Desnos M, Courbon D, Ducimetiere P. Heart-rate profile exercise as a predictor of sudden death. N Engl J Med. 2005; 352:1951–1958.

4. Kolloch R, Legler UF, Champion A, et al. Impact of resting heart rate on outcomes in hypertensive patients with coronary artery disease: findings from the INternational VErapamil-SR/trandolapril STudy (INVEST). Eur Heart J. 2008; 29:1327–1334.

5. Paul L, Hastie CE, Li WS, et al. Resting Heart Rate Pattern During Follow-Up and Mortality in Hypertensive Patients. Hypertension. 2010; 55:567–574.

6. Copie X, Hnatkova K, Staunton A, Fei L, Camm AJ, Malik M. Predictive power of increased heart rate versus depressed left ventricular ejection fraction and heart rate variability for risk stratification after myocardial infarction. Results of a two-year follow-up study. J Am Coll Cardiol. 1996; 27:270–276.

7. Diaz A, Bourassa MG, Guertin MC, Tardif JC. Long-term prognostic value of resting heart rate in patients with suspected or proven coronary artery disease. Eur Heart J. 2005; 26:967–974.

8. Caetano, J. & Delgado Alves, J. Heart rate and cardiovascular protection. Eur. J. Intern. Med. 26, 217–222, https://doi.org/10.1016/j.ejim.2015.02.009 (2015).

9. Alhalabi, L. et al. Relation of Higher Resting Heart Rate to Risk of Cardiovascular Versus Noncardiovascular Death. The American journal of cardiology 119, 1003–1007, https://doi.org/10.1016/j.amjcard.2016.11.059 (2017).

10. ACSM. ACSM Guidelines for exercise testing, 2018.

11. Wallen MP, Gomersall SR, Keating SE, Wisloff U, Coombes JS. Accuracy of Heart Rate Watches: Implications for Weight Management. PloS one. 2016;11(5):e0154420. Pmid:27232714

12. Shcherbina A, Mattsson CM, Waggott D, Salisbury H, Christle JW, Hastie T, et al. Accuracy in Wrist-Worn, Sensor-Based Measurements of Heart Rate and Energy Expenditure in a Diverse Cohort. J Pers Med. 2017;7(2). Pmid:28538708

13. Haghayegh, S, et al. Accuracy of PurePulse photoplethysmography technology of Fitbit Charge 2 for assessment of heart rate during sleep. Chronobiology International 36.7 (2019): 927–933.

14. Heneghan, C, Venkatraman S, and Russell A. Investigation of an estimate of daily resting heart rate using a consumer wearable device. medRxiv (2019): 19008771.

15. Quer G, Gouda P, Galarnyk M, Topol EJ, Steinhubl SR (2020) Inter-and intraindividual variability in daily resting heart rate and its associations with age, sex, sleep, BMI, and time of year: Retrospective, longitudinal cohort study of 92,457 adults. PLOS ONE 15(2): e0227709. https://doi.org/10.1371/journal.pone.0227709

16. Booth, M.L. (2000). Assessment of Physical Activity: An International Perspective. Research Quarterly for Exercise and Sport, 71 (2): s114–20.

17. http://www.ipaq.ki.se

18. http://www.fitabase.com

19. Chow, Hsueh-Wen, and Chao-Ching Yang. “Accuracy of Optical Heart Rate Sensing Technology in Wearable Fitness Trackers for Young and Older Adults: Validation and Comparison Study.” JMIR mHealth and uHealth 8.4 (2020): e14707.

20. Palatini et al, Predictive Value Of Night-time Heart Rate For Cardiovascular Events In Hypertension. The Abp International Study. Int J Cardiol. 2013 September 30; 168(2)

21. Ben-Dov IZ, Kark JD, Ben-Ishay D, Mekler J, Ben-Arie L, Bursztyn M. Blunted heart rate dip during sleep and all-cause mortality. Arch Intern Med. 2007; 167:2116–2121.

22. Palatini P, Thijs L, Staessen JA, et al. Predictive Value of Clinic and Ambulatory Heart Rate for Mortality in Elderly Subjects With Systolic Hypertension. Arch Intern Med. 2002;162(20):2313–2321

23. Scheer FA, van Doornen LJ, Buijs RM. Light and diurnal cycle affect human heart rate: possible role for the circadian pacemaker. J Biol Rhythms. 1999 Jun;14(3):202–12. doi: 10.1177/074873099129000614. PMID: 10452332.

24. Malpas SC, Purdie GL. Circadian variation of heart rate variability. Cardiovasc Res. 1990 Mar;24(3):210–3. doi: 10.1093/cvr/24.3.210. PMID: 2346954.

